# Sexual Dimorphism in Telomere Length in Childhood Autism

**DOI:** 10.1101/2020.04.30.20074765

**Authors:** Yasin Panahi, Fahimeh Salasar Moghaddam, Khadijeh Babaei, Mohammad Eftekhar, Reza Shervin Badv, Mohammad Reza Eskandari, Mohammad Vafaee-Shahi, Hamid Pezeshk, Mehrdad Pedram

## Abstract

Autism spectrum disorders (ASD) are lifelong heterogeneous set of neurodevelopmental conditions with strikingly profound male prevalence. Differences in sex biology and hormones are thought to play key roles in ASD prevalence and outcome, but the underlying molecular mechanisms responsible for ASD sex-differential risk are not well understood. Two recent studies reported a significant association between shortened telomere length (TL) and autistic children. However, the role of gender bias has been overlooked. Here, we carefully examined the status of average TL among nonsyndromic male and female children with autism, and we also took a close look at the data from earlier reports. A total of 58 children were recruited for this project, including 24 apparently nonsyndromic autistic children (14 males and 10 females), their healthy siblings (n = 10), and 24 sex-, age, and location-matched healthy controls. Relative TLs (RTL) were assessed by the monochrom multiplex quantitative polymerase chain reaction (MMQPCR) technique, using genomic DNA extracted from saliva samples. Data analysis showed that gender and age had strong impacts on average RTLs among the study groups. In a sex stratified manner, autistic male children had significantly shorter average RTL than their female counterparts. Only male children with autism showed a homogeneous pattern of shorter RTLs compared with their respective healthy controls. Our findings are indicative of a sexually dimorphic pattern of TL in childhood autism. The data presented here have important implications for the role of telomere biology in the molecular mechanisms responsible for ASD male bias prevalence and etiology.

## INTRODUCTION

Childhood autism, the most severe subset of autism spectrum disorders (ASD), is generally diagnosed between the ages of 2 and 9 (1) with a striking male prevalence (2). The commonly cited 4:1 male to female ratio in ASD has come under some scrutiny in recent years. However, this has not shaken the consensus on the ASD male prevalence across different cultures and regions of the world spanning several decades of epidemiological studies (3). Despite considerable advances in ASD research during the past two decades and identification of hundreds of ASD risk genes and genetic variations (4), the underlying molecular mechanisms responsible for childhood autism and its profound sex-biased risk are not well understood.

Genetic components are highly critical and play a major role (~80%) in the etiology of autism, with high heritability estimates of 64-91% based on numerous twin and family studies (5). The scope of genetic heterogeneity in autistic cases is enormous even when the portion of syndromic cases (~10%) is put aside. Yet, some aspects of autism are common and go beyond ethnic, cultural, and geographical boundaries: similar cognitive and behavioral phenotypes; profound male-biased prevalence; and shared pathophysiology, especially high levels of oxidative stress, possibly mitochondrial dysfunction, and immune/neuroimmune dysregulation and inflammation (6–8). Taken together, these features are strongly suggestive of involvement of converging pathways/processes and also sex-differential biology in ASD. A good number of ASD risk genes relate to brain development and function (in particular, synapse formation and function), and brain metabolism. However, there is also a noticeable enrichment for the genes involved in transcription regulation and chromatin remodeling (9). Although some ASD risk genes and variations are located on the sex chromosomes, they comprise only a very small portion and surely do not explain the male preponderance in autism. Overall, ASD risk genes are neither distributed nor expressed differentially in males and females. Instead, the risk genes appear to interact with and/or are modulated by sexually dimorphic processes that are associated with ASD pathophysiology, including neuron-glial interactions in the cerebral cortex. Most notably, genes related to astrocyte and microglia, which are both involved in synapse formation and function and are also upregulated in autistic brain, are expressed at significantly higher levels in the male brain (10).

Aside from hundreds of different ASD risk genes and variations distributed interstitially throughout various chromosomes, the specialized heterochromatin entities/protective caps at every chromosome ends (*i.e*., telomeres) have the potential to play some pivotal role/s in the etiology of autism. Telomeres are nucleoprotein structures composed of long stretches of 5’-TTAGGG-3’ repeats that are looped as large lariats at chromosome ends. These tightly packaged yet dynamic structures are held by members of the shelterin complex and also well-organized nucleosome particles marked with specific histone codes. Telomeres are elongated with the reverse transcription enzyme telomerase, and telomere length (TL) plays an essential role in maintaining its structure and function as it recruits the required concentration of the telomeric proteins and factors to the chromosome ends (11). Telomeres shorten with cell division and are heterogeneous in length as both the initial average TLs and the rate of telomere attrition are determined by cell type, genetic predispositions, sex hormones, and environmental factors (11–13).

In contrast to their classical characterization as barren constitutive heterochromatin, telomeres have low but important transcription activity resulting in non-coding telomere repeat containing RNA molecules known as TERRA (14, 15). Telomere length and structure impact TERRA expression levels (16) as well as transcriptional activity of subtelomeric region, an epigenetic phenomenon known as telomere position effect or TPE (17). Many telomeric factors (including telomerase itself and TERRA) have a wide range of extra-telomeric targets and functions. Telomere shortening not only impacts TPE and TERRA levels but also leads to dispersion of telomeric factors to chromosome interstitial locations affecting genome-wide transcription regulation encompassing genes involved in stress responses, metabolism, immunity, differentiation, and also the nervous system (18). It is also important to note that while short telomeres negatively impact neuronal differentiation and lead to the loss of neurons, they could enhance differentiation of glial progenitor cells and astrocytes (13).

Abnormally short leukocyte TLs have been associated with a number of psychiatric and neurological disorders and most recently, childhood autism (13, 19–21). While Li *et al*. provided the original report on shorter telomeres in autistic children using a male skewed sample from the Chinese population, Nelson and colleagues later reported a similar observation investigating a sex-skewed group of mostly White subjects from the United States. They also expanded the association of shorter telomeres to the ASD family members. Here, by recruiting comparable numbers of male and female children with moderate-severe non-syndromic autism from independent families and sex-, age-, and location-matched healthy controls from the Iranian population, as well as including the siblings of the proband cases, we take a careful look at the average TLs among males vs. females.

## RESULTS

### Demographics and Characteristics of Subjects and Study Groups

Detailed classification and demographics of all study subjects as well as the ADI-R scores of the autistic children and their verbal status are presented in the Supplementary Tables S1 and S2. Except for the sibling group, the autism case and healthy control subjects were both sex- and age-matched (Table 1). It is important to note that all of the autistic children are from independent and apparently simplex families and are within the moderate-severe bracket of the symptoms severity. Moreover, as mentioned in the Materials and Methods, all case and control subject pairs were location-matched for selection, and time-matched for sampling. It should also be noted that although six out of 10 female children with autism were not strictly speaking drug-naïve, they had extended wash out periods. As evident in Table 1, there were no significant differences in either the mean parental ages or birth weights between the study groups (all *P*-values > 0.05). Furthermore, with the exception of the siblings (8 M:2 F), in contrast to previous studies related to TL and autism, (20, 21) gender distribution was not significantly different between the cases with childhood autism and the healthy controls (*p* = 0.441).

**Table 1.** Characteristics of idiopathic autistic cases, healthy siblings, and healthy neighboring-controls.

| Variable | Case<br>(n=24) | Control<br>(n=24) | Sibling<br>(n=10) | F-value | P- value |
| --- | --- | --- | --- | --- | --- |
| Mean Age (mo) | 61.50 ±27.65 | 65.25 ±28.58 | 80.40 ±55.44 | 1.103 | 0.339 |
| Gender |  |  |  |  | 0.441 |
| Male (%) | 14 (58.33) | 14 (58.33) | 8 (80) | - | - |
| Medication (no/yes) | 14/0 | 14/0 | 8/0 |  |  |
| Female (%) | 10 (41.67) | 10 (41.67) | 2 (20) | - | - |
| Medication (no/yes) | 4/6 | 10/0 | 2/0 |  |  |
| Paternal Age (yr) | 32.9 ±5.1 | 32.3 ±4.8 | 34.8 ±4.6 | 0.970 | 0.385 |
| Maternal Age (yr) | 27.2 ±4.5 | 27.9 ±4.2 | 28.8 ±2.8 | 0.278 | 0.758 |
| Birth Weight (kg) | 3.21 ±0.50 | 3.13 ±0.41 | 3.5 ±0.64 | 1.55 | 0.220 |

**Notes:** To compare the subclinical groups, the chi-square test and a one-way analysis of variance (ANOVA) were used for gender (categorical variable) distributions and continuous variables (ages of samples, parental ages, and birth weights), respectively. The *P*-value for the gender row represents the gender distribution within the patient and healthy control groups. Medication indicates subjects had received treatment with anti-psychotic drugs for some periods in the past. The values for age and birth weight in each group are presented as mean ±SD.

### Significantly shorter avearge telomere length is evident in male children with autism but not in females

Comparison of the average relative telomere length (RTL; mean T/S ratio) values between different sample groups showed that children with autism had shorter TLs on average than both the healthy controls and apparently healthy siblings (1.09 ±0.41 vs. 1.23 ±0.47 and 1.17 ±0.40, respectively) (Fig. 1A, left), consistent with two recent reports associating telomere attrition with childhood autism and children with ASD (20, 21). After adjusting for covariates including sample’s age, sex, parental ages, and birth weight, however, these differences in average TLs were not statistically significant (β = -0.203, *p* = 0.136; and β = -0.174, *p* = 0.222, respectively). Furthermore, our analysis showed that two of the covariates had strong impacts on TL: gender (β = 0.397, *p* = 0.009) and age (β = -0.382, *p* = 0.011). Among the remaining covariates considered in the above analyses and clinical features noted in Table 1, also the maternal age for the male samples showed a noticeable association with RTL (β = 0.459, *p* = 0.027). It is also interesting to note that taking off the paired siblings, which comprise an important clinical group based on shared genetic background and environment with their respective index patients but are neither age- nor sex-matched with the autism and control groups, does not change the outcome of autistic vs. the control RTL analysis significantly (β = -0.222, *p* = 0.090). However, it does highlight a significant association between birth weight and RTL values (β = 0.348, *p* = 0.011).

**Figure 1.**
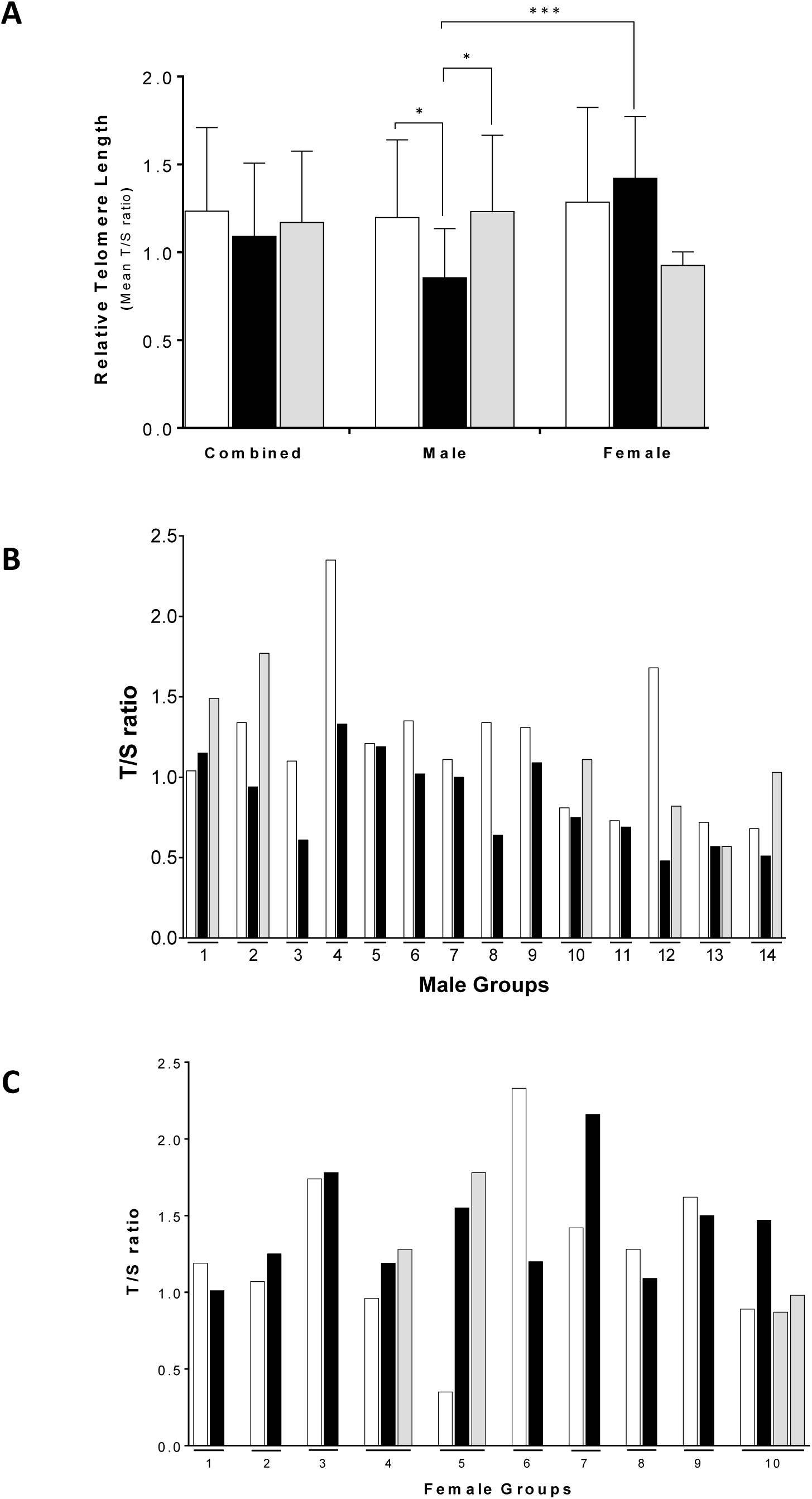
Average relative telomere lengths (RTLs; T/S ratios) for different clinical groups and individually matched groups. Autistic subjects (in the middle), healthy siblings (to the right), and close neighboring healthy controls (to the left) are shown with black, gray, and white bars, respectively. **(A)** Comparison of the total average T/S ratios (mean ± SD) in children with idiopathic autism, their healthy siblings, and healthy neighboring controls in both sexes (combined), and in male and female subjects exclusively. **(B and C)** Comparison of the individual T/S ratios for patients, siblings, and healthy neighboring controls in male **(B)** and female **(C)** autism individually matched groups. *, *p* < 0.05; ***, *p* < 0.001.

Interestingly, organizing different groups based on gender revealed a strikingly different picture on RTL between the male and female children with autism. The scatter plots of the individual average T/S ratios for all samples organized by various subdivisions are presented in the Supplementary S1. The female children with autism had a significantly longer mean RTL when compared with their male counterparts (1.42 ±0.35 vs. 0.85 ±0.28; β = 0.792, *p* = 0.0002) (Fig. 1A, middle and right). In fact, the male children with autism had a significantly shorter RTL compared with their sex-matched respective healthy controls (0.85 ±0.28 vs. 1.19 ±0.44; β = -0.374, *p* = 0.028). The male children with autism also had a noticeably lower mean T/S ratio than their siblings (0.85 ±0.28 vs. 1.23 ±0.43; β = -0.369, *p* = 0.030). By contrast, the situation was almost reversed among females: the female patients with autism showed even a higher mean T/S ratio compared with either their matched healthy control group (1.42 ±0.35 vs. 1.28 ±0.53) or siblings (1.42 ±0.35 vs. 0.92 ±0.07). However, these differences are not statistically-speaking significant (β = 0.178, *p* = 0.533; and β = 0.575, *p* = 0.147, respectively), especially the latter because of the very small number of female siblings (N=2) in our study. Finally, we found no significant correlations between clinical evaluations/ADI-R total scores and RTL values within different patient groups (Supplementary Material, Fig. S2), except for seemingly interesting contrasting trends for male vs. female nonverbal autistic children (panel E).

### Sexually-dimorphic patterns of RTL among individuals with childhood autism

Intriguing patterns and contrast emerge when children with non-syndromic autism are divided based on gender and the T/S ratios are presented/organized by individually-matched subject groups (*i.e*., each child with autism paired with his/her respective sex-, age, and location-matched healthy control, plus the proband’s sibling if available) (Fig. 1B and 1C; and Supplementary Material, Table S2). As shown in Fig. 1B, all of the male autistic subjects had shorter RTLs than their matched healthy controls except for one, male sample group no. 1, in which the control child was 27 months older than the child with autism. It is also noteworthy that in sample group no. 13, the male child with autism had an RTL that was almost equal to his healthy brother, while the proband was more than 3.5 years younger than his sibling. In contrast to the pattern described for male children, distribution of the T/S ratios among the female individually-matched groups appear heterogeneous and does not seem to follow a clear pattern (Fig. 1C; and Supplementary Material, Table S2). While six out of 10 female children with autism had longer average TLs than their individually-matched healthy controls, the remaining four had shorter telomeres. It is not clear if medication exposure (female patients no. 1, 4, 5, 6, 8, and 10) had any impact on TL because of the small number of such samples in this study. Not to mention that three out of the four drug-naïve female patients (no. 2, 3, and 7 vs. no. 9) also had longer telomeres than their respective controls (Fig. 1C).

### The rate of telomere attrition is higher in children with autism

Overall, the inverse correlation between individual T/S ratios and the age of the samples was indicative of telomere attrition with increase in age (Fig. 2A). The biggest contribution to telomere shortening with age, considering Pearson’s correlation coefficient (*r*) values, came from children with autism (Fig. 2B and 2D). However, in contrast to the differential average RTLs between male and female children with autism, it did not appear that the telomere attrition rate was sexually biased among autistic children (male: *r* = -0.50, *p* = 0.06; female: *r* = -0.51, *p* = 0.15, respectively). Finally, the telomere attrition rate among the healthy siblings (*r* = -0.47, *p* = 0.16) appeared to be about the same as the children with autism and higher than the healthy male and female controls (Fig. 2C, 2E, and 2F). However, these differences were not statistically significant, which was likely because of having a limited number of samples in each group.

**Figure 2.**
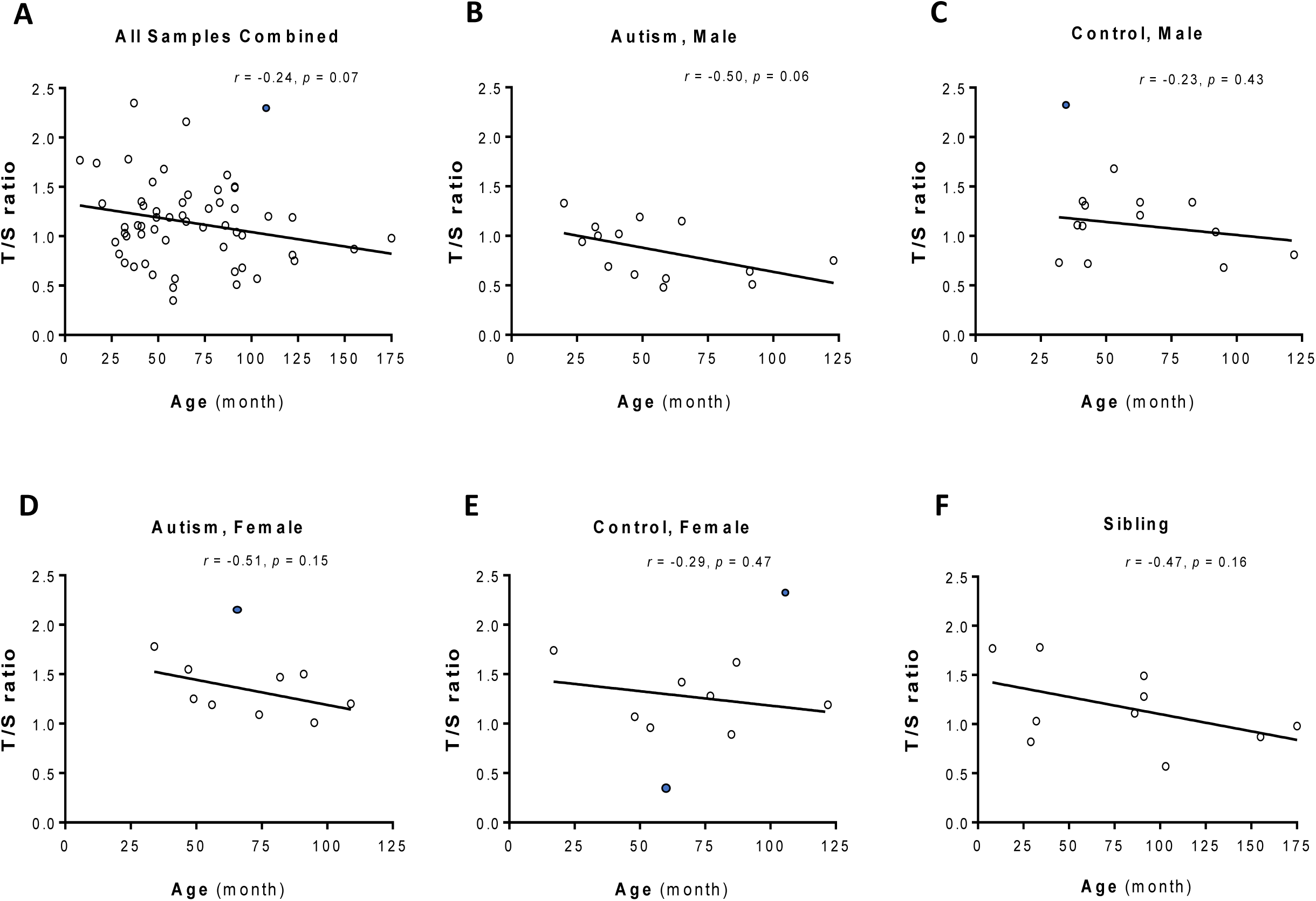
Telomere attrition with age. Correlation between decreasing average RTLs (T/S ratios) and age in all subjects combined **(A)**, male patients **(B)**, male healthy controls **(C)**, female patients **(D)**, female healthy controls **(E)**, and siblings **(F)**. Pearson’s correlation coefficient (*r*) values indicate how much the age and T/S ratios in subclinical groups are interconnected. The filled circles represent the outlier data points.

## DISCUSSION

Abnormal telomere attrition in leukocytes has been linked to a number of human diseases and psychological disorders including two recent reports on autistic children (20, 21). Our investigation and the findings presented here show that the abnormal reduction of TL in childhood autism follows a sexually dimorphic pattern. The male children with autism followed a surprisingly homogeneous pattern of shorter telomeres compared with their sex-, age-, and location-matched healthy controls. By contrast, the female cases with autism not only did not follow such a clear pattern, but also as a group exhibited no evidence of shorter telomeres compared with their matched healthy control group. In fact, six out of the total of 10 female children with autism had considerably longer average TLs than their individually-matched controls. Furthermore, there was a clear difference in TL between the male and female groups of children with autism, with the female children having a significantly longer average RTL.

At first look, our data showing a male bias for telomere attrition in childhood autism may seem contradictory to the recent reports by Li *et al*. (20) and Nelson *et al*. (21) However, a careful examination of the data from these studies reveals an intriguingly different picture. A major strength of our study is focusing on a well-defined, yet highly important clinical subset of children with autism. The clinical homogeneity of the autistic patients in the present study is significantly higher than the two previous reports. It is not clear if in the study by Li *et al*. they have recruited some of their patients from high functioning category or multiplex families. However, from the mean CARS (<u>C</u>hildhood <u>A</u>utism <u>R</u>ating <u>S</u>cale) scores presented in the report (34.61 ±3.09), it is quite evident that the autistic children belong to two brackets of symptoms severity, with most of them coming from the clinically “mild-moderate” bracket (*i.e*., CARS score of 30.0–36.5) (20). As for the study by Nelson and colleagues, they clearly note that their autistic families (which they call HRA for high-risk for autism), had “at least” one child with “community diagnosis of ASD” (21). This indicates that the autistic children were recruited based on a wider definition of ADS and some from multiplex families (*i.e*., multiple affected children that are typically higher functioning with mild-moderate clinical symptoms). The authors do not provide any detailed information on the severity of symptoms or distribution of the autistic children from multiplex families in the paper (21).

Another major strength and difference is that the autistic children and their healthy controls were carefully sex-, age-, and location-matched in the present study. As presented in the Supplementary Table S3, in the study by Li *et al*., in a clear contrast to our study, the gender distribution was highly skewed in favor of the male subjects, particularly among the children with autism (98 M:12 F and 98 M:31 F in patient and healthy control groups, respectively). Despite having a disproportional number of the healthy controls (N=31) vs. the number of female children with autism (N=12), Li and colleagues report that they found no significant difference in RTL between their female patients and the controls (1.01 ±0.44 vs. 1.06 ±0.38, *p* = 0.734). The authors also state that they saw no significant difference in TL between their male and female subjects in general (*i.e*., combined groups) (20)^(p2)^. Apart from a statistical argument, we find it surprising that the authors simply disregard the noticeable size difference (17%) in the RTL values between their male and female cases with autism (0.86 ±0.25 vs. 1.01 ±0.44, respectively). Interestingly, this difference was even slightly larger than the difference reported between their male and female combined cases with autism and the controls (0.88 ±0.28 vs. 1.01 ±0.43, respectively). We should point out that a 17% increase in T/S ratio from male to female autism cases could translate to hundreds of base pairs in TL difference. As for the study by Nelson and colleagues, there were only two female children included among the total of 18 cases with autism presented in the study. Surprisingly, the RTL values between the two female cases with autism and that of the 16 male cases were markedly different (0.50 ±0.01 vs. 0.70 ±0.24, respectively). However, the ages of the two female children with autism were not disclosed and it was not clear if they came from independent families. Furthermore, because of the very skewed male to female ratio (16 M:2 F) in the autism group, it is extremely difficult to draw any conclusions regarding a meaningful association between TL and gender among the patients. It is also interesting to note that in their younger than ASD proband sibling/infant group that included 16 male vs. 12 female children, whom Nelson *et al*. appropriately characterize as high-risk cases for autism, the average RTL in the female subgroup was noticeably longer than their male counterparts (0.87 ±0.15 vs. 0.73 ±0.46, respectively) (21).

The relationship between T/S ratios and age among our study groups are in line with a number of highly cited previous studies showing a strong “negative correlation” between age and TL in leukocytes (22–24). Not only did we see a clear trend of telomere shortening with increase in age among the entire spectrum of subjects in our study (Fig. 2A), but also apparently a higher rate of reduction in TL among the cases with autism, followed closely by the siblings, in comparison to the healthy controls (Fig. 2B-F). Consistent with our findings, Nelson *et al*. reported an inverse correlation between age and TL in their entire sample (*r* = -0.448, *p* < 0.001). However, the authors do not elaborate on the status of their different clinical groups [see (21)^(p591)^ the 1^st^ paragraph]. By contrast, while Li *et al*. found a significant inverse correlation between TL and age in their controls (*r* = -0.30, *p* = 0.0001), surprisingly, they reported a positive correlation between TL and age across both their entire sample and within their autism cases (*r* = 0.24, *p* = 0.001; and *r* = 0.178, *p* = 0.064, respectively) (20)^(p2)^. Later in their Discussion section, the authors attribute the unexpected positive association of TL and age among their autism cases to the patients already having shorter than normal telomeres as a possible reason (20)^(p4)^. This is in sharp contrast to our observations. In addition to the possibility of the difference in the study design, sampling source and method as possible causes for the discrepancies regarding RTL and age in the study by Li *et al*., another factor could be a difference in the efficiency of the different qPCR methods used in these studies. The improved MMQPCR technique used in the present study gives a considerably more accurate T/S ratio for each sample (see Supplementary Material Text S1 for a more detailed discussion).

There have been some interesting discussions presented on the potential role of telomeres and in particular shortened TL in childhood autism. Li *et al*. have put forward three different scenarios: 1) cause, 2) effect or 3) “unrelated” but originating from shared mechanisms. Although they leave open the possibility of telomeres being involved in the molecular mechanism and pathology of autism, the authors seem to favor the third scenario as they found no correlation between TL and clinical parameters (20). Nelson and colleagues on the other hand make a convincing argument by citing a number of earlier studies in the literature implicating the possibility of telomere structure and function in ASD susceptibility. The authors conclude that not only shorter TL has to be taken as an indicator of both genetic and environmental liabilities but also a serious risk factor for future physical and psychological complications (21). We find the argument presented by Nelson *et al*. to be more plausible with one important note: shorter than normal TL seems to be applicable only to boys with childhood autism not girls. When combining both sexes, it would be more proper to say “abnormal” telomere length/s and/or perhaps structure/s, especially at the individual level. Telomeres are not neutral chromatin entities, and taking into account the significant roles which telomeres and their associated factors play in a wide range of important cellular processes (encompassing the nervous system) (18, 25), it is hard to imagine abnormal telomere length/s not playing any roles in the etiology of childhood autism and the underlying molecular mechanisms involved.

The evidence/argument we provide here, supporting a male bias for abnormally shortened telomeres in children with autism, provides a link between RTL and sex biology and the striking 4:1 male prevalence in autism. Telomeres could play a pivotal role in connecting and explaining the molecular mechanisms underlying the “multi-factorial sex/gender-differential liability model” of autism, with interplay of female protective factors (e.g. estrogen exposure and homogametic sex/X chromosomes) and male-specific risk factors (e.g. pre-/perinatal fetal testosterone, not having a paternal X chromosome, and unguarded Y chromosome) (3, 26).

There is a little doubt that excessive oxidative stress plays a key role in the etiology of autism (6, 27). The sexually dimorphic patterns of TL in autism could be explained by interplay of reactive oxygen species (ROS), sex hormones, and telomere-length homeostasis (12, 28). Excessive amounts of ROS cause rapid telomere shortening (29) due to DNA breaks (30). Sex hormones can modulate the impact of ROS on telomeres: while the female primary sex hormone, estrogen, can protect against telomere shortening, testosterone, the male primary sex hormone, can exacerbate telomere erosion. There are compelling lines of evidence that estrogen prevents telomere shortening through antioxidant activities and also activation of telomerase *via* a number of mechanisms including suppression of free radical production, sequestration of ROS, and modulation of antioxidant enzyme activities (12). Furthermore, ovarian steroid hormones, and in particular estrogen, have both neurotrophic and neuroprotective functions (31).

In support of the above arguments, long time estrogen therapy lead to attenuation of telomere erosion in post-menopausal women through antioxidant mechanisms (32). Also longer telomeres in female than male adults have been attributed in part to a slower rate of telomere attrition during early years of female life possibly due to estrogen effect (33, 34). In a recent study by Windham *et al*., it has been shown that lower levels of maternal unconjugated/free estriol, which is a circulating active form of estrogen, during mid-pregnancy, significantly increases the risk of ASD in children (35).

In contrast to the protective effects of estrogen, the primary male sex hormone testosterone has been linked to increased susceptibility to oxidative stress (36, 37) and shorter TL in youth (38). Baron-Cohen and colleagues, have reported a strong association between increased levels of fetal testosterone and children later being diagnosed with ASD (39). Although difficult, it would be highly informative to find out how the status of blood or saliva leukocyte TL in autistic children relates to the actual status of TL in brain cells, and how that in turn affects neuron-glial interactions. In an in-depth analysis of available post-mortem brain cortex data sets, Werling and colleagues have shown that although ASD risk genes appear to be distributed non-differentially in both sexes, they seem to interact with sexually dimorphic pathways associated with ASD pathophysiology. The investigators found a high enrichment for gene expression related to microglia, the residing innate immune cells in the brain, and astrocytes in ASD cortex. These genes are typically expressed at higher levels in the male brain. By contrast, the genes that are typically expressed at higher levels in the female brain (most notably genes related to synapse formation and function) were found at depleted expression levels in autistic cortex (10). The basis of sex-differential neuroanatomy are established very early in life during a critical period of brain development, a process mainly driven by the primary male steroid hormone, testosterone (40). This so-called “masculinization” of the brain also involves inflammation signaling molecules and neuroimmune system (in particular, activated microglia and reactive astrocytes), prompting a proposed model for convergence of masculinization of the brain, inflammation, and neuroimmune system in the male-biased vulnerability to ASD (7). The details of the molecular and cellular mechanisms involved are poorly understood. Considering the evidence on TL and telomerase expression levels differentially affecting neuronal vs. glial differentiation, (13) sexual dimorphism of TL could impact, and/or act as a biomarker of, such processes in childhood autism.

An important point to highlight here is the fact that similar to the works by Li *et al*. and Nelson *et al*., in the present study, we have investigated the average RTL for all 46 pairs of human chromosomes combined (*i.e*., an average relative length for a total of 92 telomeres combined). It remains to be seen what the status of different chromosome-specific telomeres are in autism cases. Although under normal circumstances, TL is an important factor in telomere structure, it is not necessarily a conclusive factor (41). Our own investigation indicates that although female children with autism have significantly longer average RTLs than their male counterparts, they also show significantly high concentrations of oxidized bases, which is an indication of oxidative damage, at their telomeres. Furthermore, despite the fact that autistic male children have significantly shorter average RTLs than the healthy controls, they show lower levels of TERRA transcription from certain chromosome ends (M. Eftekhar and Y. Panahi *et al*., manuscript in preparation).

Despite some important differences, the fact that three independent studies on three geographically, ethnically and culturally diverse populations find strong associations between TL dynamics and childhood autism is highly significant. It further highlights the importance of genetic factors and suggests that telomeres are important players in the etiology of autism. Our observations on the sexually dimorphic patterns of TL in children with non-syndromic autism could provide important insights for deciphering telomere dynamics and the underlying molecular mechanisms of childhood autism. They also open the door for some important investigative questions: Do sexually dimorphic average TLs reflect the global status of TLs in males and females? What is the status of telomere structure and oxidation in children with autism? What is the status of telomeric transcription in children with autism? Could TL status (abnormally shorter or longer) be taken as a biomarker for diagnosis, further investigation, and/or treatment in autism? Answering these questions will provide additional clues for a better understanding of the role/s of telomere biology in autism.

## MATERIALS AND METHODS

### Subjects and Sampling Procedure

A total of 58 participants were recruited for this study, including 24 children (14 males and 10 females) diagnosed with idiopathic (i.e., non-syndromic) autism (61.50 ±27.65 months), 10 healthy siblings (80.40 ±55.44 months), and 24 age-, gender-, and location-matched typically developing children as non-family healthy controls (65.25 ±28.58 months). All cases were collected during a three-year period from 10 different cities spanning nine provinces. Children with idiopathic autism were diagnosed as previously described (42). Briefly, each child with autism was diagnosed by at least two qualified clinicians, a child neurologist and a specialized psychiatrist, according to the Diagnostic and Statistical Manual of Mental Disorders fourth edition, Text Revised (DSM-IV-TR) criteria. Diagnosis of autism was confirmed by Autism Diagnostic Interview-Revised (ADI-R), a standardized semi-structured diagnostic algorithm for autism based on the definitions set by DSM-IV and International Statistical Classification of Diseases and Related Health Problems 10th Revision (ICD-10) (43, 44). Autistic subjects diagnosed with any other medical disorders and/or genetic syndromes were not included in the study. Autistic cases from multiplex families (*i.e*., having more than one affected child), high functioning autism category, and “mild-mederate” severity of symptoms clinical bracket were not included in the study. Sex- and age-matched healthy control children were picked from the same vicinity or close neighboring locations as their respective probands, and they were screened by the Strengths and Difficulties Questionnaire (SDQ) (45). Male patients were newly diagnosed and were antipsychotic-drug naïve subjects during sampling. In contrast, because of the limited number of female children with non-syndromic autism available, only four out of 10 female patients were truly drug naïve. As a cautionary measure, when an equal-age matched qualified control was not available in the close vicinity of a proband, when given the choice, preference was given to slightly older controls over those younger than the respective proband. The study had the approval of the ZUMS Research Ethics Committee (ZUMS.REC.1392.97), it was conducted in accordance with the Declaration of Helsinki, and all children had signed informed consent provided by their parents or caregivers.

Saliva sampling was done using the Oragene OG-250 collection kits (DNA genoTeK Inc., Ottawa, Ontario, Canada) following manufacturer’s protocol.

### Genomic DNA Extraction and Preparation

Genomic DNA (gDNA) was extracted from whole saliva samples, using the manufacturer’s protocol (OG-250; DNA genoTeK Inc., Ottawa, Ontario, Canada) with slight modification. Briefly, prior to DNA extraction, each sample was incubated at 50°C for 1 hour using a water bath. Following this step, a 500-µL aliquot from the sample was transferred into a 1.5-mL microcentrifuge tube for processing and the remaining sample was stored at -20°C for potential use in the future. Twenty µL of the Oragene DNA purifier solution (PT-L2P) was added into the microcentrifuge tube containing the sample and mixed by vortexing for a few seconds. Following a 10-min incubation on ice, the sample mix was centrifuged at 15,000 xg for 10 min at RT. The supernatant was carefully transferred into a fresh microcentrifuge tube using a micorpipetter, 500 µL of RT 100% ethanol was added into the tube and mixed by inverting for 10 times. The tube was then allowed to stand at RT for 10 min to allow for full precipitation of the DNA, followed by centrifugation at 15,000 xg for 2 min at RT, followed by carefully discarding the supernatant and washing the DNA pellet with one mL of RT 70% ethanol. The air-dried DNA pellet at the final step was dissolved in 100 µL of a customized TE buffer with 1/10 of EDTA concentration (10 mM Tris, 0.1 mM EDTA, pH 8.0), followed by incubation at 50°C for 20 min in a water bath. The samples were left at 4°C overnight for dissolving gDNA homogenously, and then quantified by NanoDrop 2000c spectrophotometer (Thermo Fisher Scientific, DE, U.S.A.) and also ran on 0.8% agarose gel to check for the gDNA integrity. The A260/A280 absorbance ratio for the DNA samples typically ranged between 1.7 and 1.95. For the occasional instances with the A_260_/A_280_ absorbance ratios below 1.6 (because of high protein content), the gDNA extraction procedure was repeated. In a very few cases that the A_260_/A_280_ absorbance ratios remained suboptimal, the extraction aliquots for each case were pooled and cleaned up with Proteinase K (46).

### Telomere Length Assay; Monochrome Multiplex-qPCR

The relative average telomere lengths (RTLs) were determined employing the Monochrome Multiplex qPCR (MMQPCR) telomere length measurement assay originally developed by Richard Cawthon (47) with slight modifications. PCR amplifications were done using Real Q plus 2x Green Master Mix, high ROX (Ampliqon, Herlev, Denmark) by adding 2 mM MgCl_2_, 5% DMSO per sample. Each reaction contained 900 nM of each telomere primer (Telg: 5′-

ACACTAAGGTTTGGGTTTGGGTTTGGGTTTGGGTTAGTGT-3′ and Telc: 5′-TGTTAGGTATCCCTATCCCTATCCCTATCCCTATCCCTAACA-3′), 900 nM each albumin gene (ALB) primer (Albu: 5′-CGGCGGCGGGCGGCGCGGGCTGGGCGGaaatgctgcacagaatccttg-3′ and Albd: 5′-GCCCGGCCCGCCGCGCCCGTCCCGCCGgaaaagcatggtcgcctgtt-3′), and 20 ng of the gDNA sample in a final volume of 20 µL. In each run, a fixed human reference sample and a no-template control (NTC) were included. All reactions were run in triplicates.

Amplifications were carried out using QIAGEN Rotor-Gene Q (former Corbett Rotor-Gene 6000, Dusseldorf, Germany) qPCR platform under the conditions previously described by Cawthon (47): one cycle of 95°C for 15 min, 2 cycles of 94°C for 15 s and 49°C for 15 s, 35 cycles of 94°C for 15 s, 62°C for 10 s, 74°C for 15 s with signal acquisition for telomere, followed by 84°C for 10 s, and 88°C for 15 s with signal collection for single copy gene (ALB) and concluded with a melting curve ramping from 72°C to 95°C, rising by 0.5°C in steps of 30 s. To determine relative telomere to single-copy gene (T/S) ratios and efficiency, two standard curves were produced by serial dilution of the reference DNA (150-0.61 ng/µL, threefold, six points). The slope of the standard curve for telomere and ALB were -3.01 and -3.39, respectively. The coefficient of regression values for both standard curves were 0.99. Reaction efficiency for telomere was 1.14 and for albumin was 0.96. The average coefficient of variation (CV) within triplicates were 0.72% for telomere assay and 0.93% for ALB assay separately, and the average inter-assay CV for the T/S ratio was 7.92%.

### Statistical Analysis

All statistical tests were two-sided, with the *P*-values below 0.05 (*p* < 0.05) considered as statistically significant. Statistical analysis was performed using the SPSS software, ver. 22.0 (IBM Corp., Armonk, NY, U.S.A.). Scatter plots and bar graphs were made using GraphPad Prism, ver. 6.07 (GraphPad Software Inc. La Jolla, CA, U.S.A.). Distributions of mean RTL values among clinical groups were normal assessed with the Shapiro-Wilk tests and visual inspection of their respective histograms and plots. To compare variables between subclinical groups, chi-square test was used for gender, as a categorical variable, and a one-way analysis of variance (ANOVA) for continuous variables including age of samples, parental ages, and birth weight. Because TL may be affected by confounding factors including the age of samples, parental age, weight of birth, and gender, multiple linear regression model was used for comparing the mean RTLs between different groups with dummy variable coding. Collinearity test was used to examine potential correlations among independent variables. Since distributions of ADI-R scores were non-normal based on inspection of their respective histograms and/or Shapiro-Wilk tests, a nonparametric method, Spearman’s rho correlations were used to examine the association between RTL values and ADI-R total scores in patients.

## Data Availability

The main body of the data is provided either in the text of the manuscript or as supplementary information.

## Supplementary Material

A composite PDF document containing the following:

Three Tables (Tables S1-S3); Two Figures (Figs. S1-S2); One Text (Supplement Text S1); and References (total of 3).

## Acknowledgements

This work was supported by Zanjan University of Medical Sciences (ZUMS) grant numbers A-12-534/1-4 and A-12-534/6-8, and it had the approval of the ZUMS Research Ethics Committee (ZUMS.REC.1392.97). We would like to thank Zahra Ghasemi, Mandana HadiJafari, Karim Dadashi Noshahr, and Elham Rostami for their help in the recruitment of the patients, sample collection, identification of the locally-matched controls, and independent ADI-R scoring. We would like to thank Joseph A. Baur, Ph.D. (Dept. of Physiology, University of Pennsylvania) and John P. Murnane, Ph.D. (Dept. of Radiation Oncology, University of California, San Francisco) for careful reading of an earlier version of this manuscript and providing valuable comments. We are also thankful to Mohammad H. Rahbar, Ph.D. (Dept. of Internal Medicine, UT Health Science Center at Houston) for his helpful comments and suggestions during the 2015 IMFAR meeting in Shanghai, China.

## ABBREVIATIONS

ASD: Autism spectrum disorders
ADI-R: Autism Diagnostic Interview-Revised
ANOVA: Analysis of Variance
CV: Coefficient of Variation
DSM-IV-TR: Diagnostic and Statistical Manual of Mental Disorders, Fourth Edition, Text Revision
ICD-10: International Statistical Classification of Diseases and Related Health Problems version 10
MMQPCR: Monochrome Multiplex Quantitative PCR (**qPCR**)
RTL: Relative Telomere Length
TERRA: Telomeric Repeat Containing RNA
TL: Telomere Length
TPE: Telomere Position Effect
T/S ratio: relative telomere to single-copy gene ratio

